# A Pilot Study on Serum Lipidomic Alterations in Patients with Adrenal Tumors

**DOI:** 10.64898/2026.07.01.26356676

**Authors:** Michaela Chocholoušková, Filip Čtvrtlík, Zbyněk Tüdös, Igor Hartmann, Jan Schovánek, Jitka Vostálová, Jitka Prošková, Karel Pacak, Michal Holčapek

## Abstract

Adrenocortical carcinoma (ACC) is a rare, aggressive malignancy posing significant diagnostic challenges, particularly in distinguishing it from other adrenal tumors, such as adenoma and pheochromocytoma, due to overlapping imaging and biochemical features. Improved non-invasive tools are critically needed for earlier, more accurate classification of this rare cancer. This pilot study analyzed serum lipidomic profiles in ACC, pheochromocytoma, and adenoma patients versus healthy volunteers. The most significant alterations occurred in sphingomyelins (SM) and diacylglycerols (DG). All tumor samples showed reduced very-long odd-chain SM (e.g., SM 39:1, SM 41:1, SM 41:2) and elevated DG (e.g., DG 34:1, DG 34:2, DG 36:2). These abnormalities were most pronounced in malignant tumors: ACC and metastases (AUC = 0.933), followed by pheochromocytoma (AUC = 0.800) and adenoma (AUC = 0.711). ACC patients also exhibited specific lipid signatures with decreased alkyl/alkenyl phospholipids (e.g., PE O-38:5) and lysophosphatidylcholines (e.g., LPC 20:5, LPC 18:2) versus healthy volunteers, not observed in pheochromocytoma or adenomas. Ceramide species (e.g., Cer 42:2;O2, Cer 34:1;O2) were increased in ACC compared to the other tumor types. Incorporating lipid-to-lipid ratios (Cer/SM, Cer/DG) further improved statistical model accuracy. Compared to clinical biochemistry/oxidative stress (OS) parameters, lipidomic profiling showed superior discriminatory power in adrenal tumor diagnosis. The presented study shows the serum lipidomic profiling as a promising non-invasive method for distinguishing adrenal tumor subtypes — ACC, pheochromocytoma, and adenoma — from healthy individuals, with strong diagnostic potential for ACC.

## INTRODUCTION

The adrenal glands are vital components of the endocrine system, responsible for the production of hormones essential for homeostasis, including cortisol, aldosterone, and catecholamines. Pathological conditions affecting the adrenal glands, such as adrenal adenoma, pheochromocytoma, adrenocortical carcinoma (ACC), and adrenal metastasis, can present with diverse clinical symptoms/signs, necessitating careful and precise diagnostic evaluation [1, 2].

The differential diagnosis of adrenal tumors is often complex and requires a multidisciplinary approach that combines detailed imaging techniques, such as anatomical (*e.g.*, computed tomography and magnetic resonance imaging (MRI)) and functional (*e.g.*, positron emission tomography (PET)) imaging, together with laboratory assessments of hormonal profiles and genetic markers as well as histopathological information [3]. The imaging plays a crucial role in the detection and characterization of adrenal tumors, providing essential information about the size, morphology, and functional status of these lesions [4]. Adrenal adenomas are typically well-defined, small lesions that can be difficult to distinguish from other tumors, especially in cases of larger tumors and increased attenuation [5]. Adenomas, although functionally distinct from pheochromocytomas due to catecholamine and metanephrine production, both tumor types may exhibit overlapping imaging characteristics, particularly in case of lipid-poor adenomas or for imaging without functional assessment [6, 7]. ACC are very rare but aggressive malignancies, which can present as large and heterogeneous masses that can be difficult to differentiate from adrenal metastases, especially when metastases from primary cancers, such as lung, breast, or renal cell carcinoma mimic the appearance of primary adrenal malignancies, particularly in patients in whom primary cancer has not been diagnosed [8].

Biochemical tests are used to evaluate the hormonal activity in adrenal tumors. They play a significant role in the diagnostic process. However, conventional biochemical assays have limitations that might lead to diagnostic uncertainty [9, 10]. Adrenal adenomas may not release any hormones or may produce hormones in levels that are within normal reference ranges, resulting in false-negative or overlooked diagnoses. Similarly, biochemical tests for pheochromocytoma, such as plasma or urinary metanephrines, can sometimes yield false-negative or false-positive results, particularly in patients with either very small tumors or those on drugs or with renal failure [11, 12]. Furthermore, ACC and adrenal metastases may exhibit non-specific or inconclusive biochemical profiles, complicating their differentiation from other adrenal masses. The variability in test results can be influenced by comorbid conditions, adding another layer of complexity to the diagnostic process [10].

Despite recent significant diagnostic and imaging advances, the differentiation of various adrenal tumors including adrenal adenomas, pheochromocytomas, and ACC can be challenging due to overlapping imaging features, pitfalls, and gray zones in biochemical tests. Therefore, the use of novel non-invasive methods that can both refine the diagnostic process, while enhancing understanding of the pathophysiology of adrenal tumors is very needed.

Lipid functions in the human body include components of cellular membranes, energy storage, and signal transduction. Alterations in the lipid metabolism can affect cellular processes, such as cell growth, proliferation, and differentiation, with uncontrolled cell growth and proliferation as well as enhanced migration, which are the basic characteristics of cancer. These abnormal cellular processes make a close connection between altered lipid metabolism and cancer development and progression [13-15].

Several studies have shown that oxidative stress (OS) plays a significant role in cancer development and progression. For example, reactive oxygen species (ROS) can initiate DNA mutations, which affect genes that regulate cell growth, leading to cancer initiation and also contributing to cancer progression, invasion, migration, and metastasis [16-22].

In the present study, we evaluated the serum lipidomic profiles of patients with malignant lesions (ACC and metastases), benign adenoma and pheochromocytoma as well as healthy controls (volunteers without any adrenal pathology), particularly in order to develop a new promising diagnostic approach to ACC. Moreover, this study evaluated selected clinical biochemistry/OS parameters that could be used as additional supporting information in the diagnosis of adrenal tumors, particularly ACC.

## RESULTS

### Patients, Healthy Volunteers, and Clinical Information

The present retrospective study involved 40 healthy volunteers (controls) and 42 patients with tumors. The tumor samples were divided into 4 groups as follows (**Fig. 1A**): primary ACC, metastases (various tumors), adenoma, and pheochromocytoma. There were no significant differences between the anthropometric characterization of healthy subjects and patients. **Fig. 1B** illustrates the similar age and BMI distribution among both groups. All detailed clinical information including values from clinical biochemistry and OS parameters is summarized in Supplementary Material **Table S1.**

**Figure 1.**
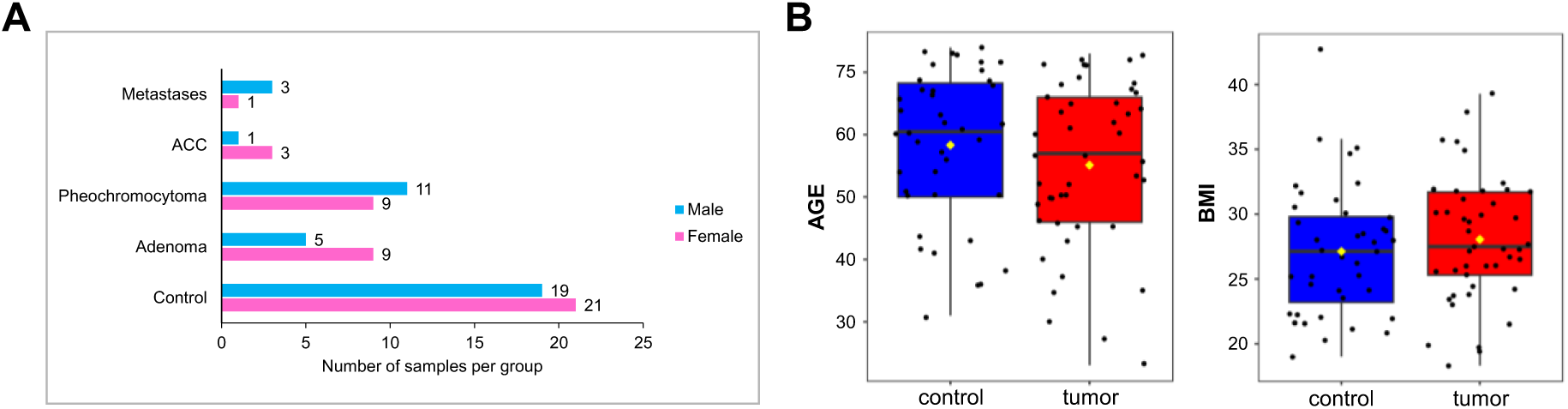
Study characteristics: (**A**) characterization of control and patient groups (the initial collection), (**B**) distribution of age and BMI between two groups, controls and patients.

### Differences Between Healthy Volunteers and Patients with Adrenal Tumors

First, we concentrated on identifying differences in serum lipidomic profiles among subjects with primary adrenal tumors, metastases and healthy subjects, specifically focusing on lipid classes that are highly abundant in serum. The principal component analysis (PCA) score plot presented in **Fig. 2A** and **2B** illustrates a partial discrimination between two groups: healthy subjects and patients with adrenal tumor subtypes – ACC, adenoma, pheochromocytoma – and metastasis, including data from both sexes. The PCA loading plot (**Fig. 2C**) highlights the contributions of lipid species and classes to the principal components. The lipid classes that contributed the most significantly to differentiation between two groups were SM, DG, and TG. SM exhibited an opposite contribution to the differentiation between these two groups in comparison to DG and TG. Box plots created for the sum of lipid species within SM, DG, and TG lipid classes are presented in **Fig. 2D**. Overall, the levels of SM were decreased in patients with primary adrenal tumors and metastasis while DG and TG levels were increased. To gain deeper insights into the behavior of specific molecular species within each lipid class or subclass, we further analyzed individual lipids and compared their concentrations between tumor and control samples.

**Figure 2.**
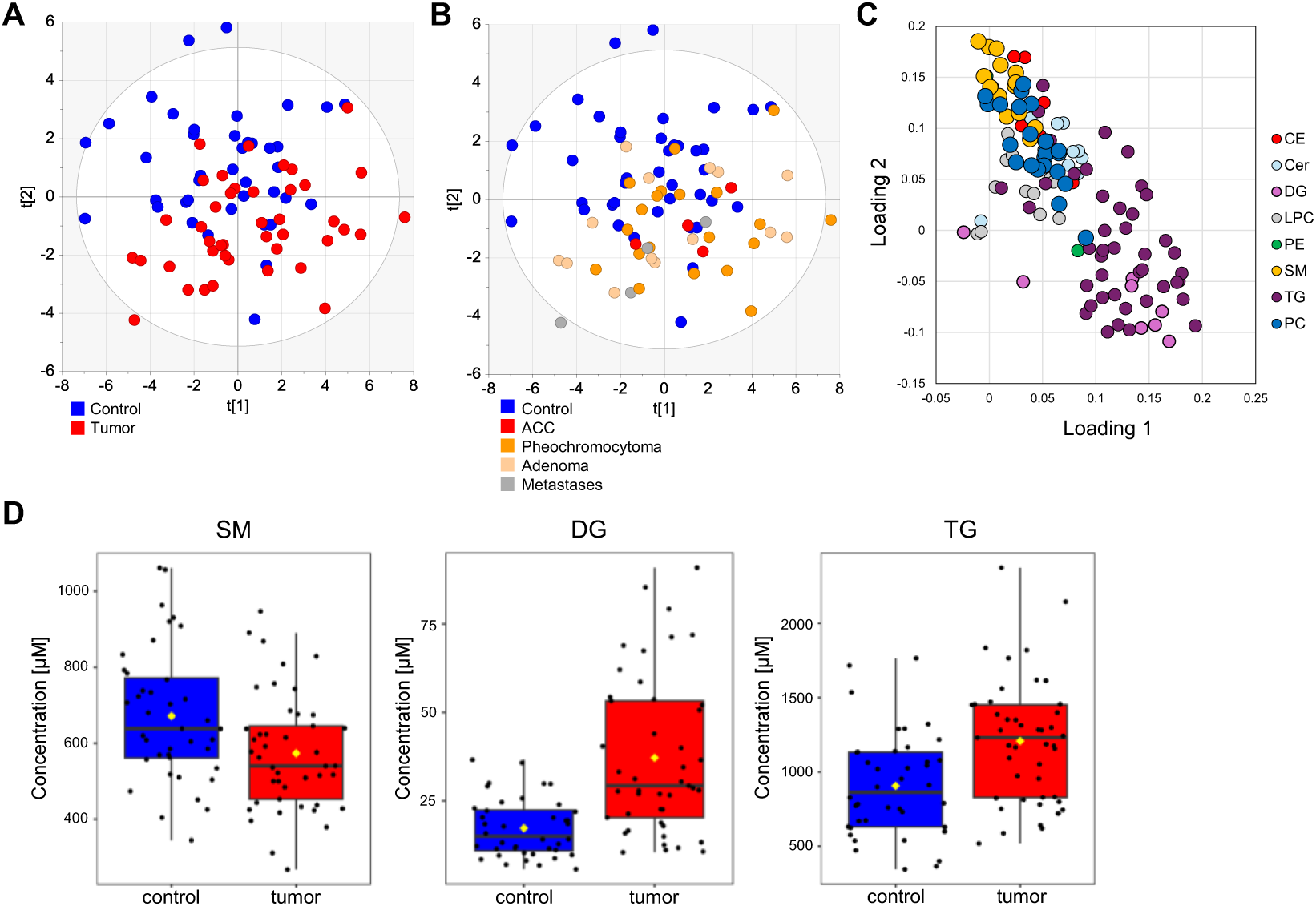
Basic characteristics of the lipid dataset for patients with adrenal tumors and healthy subjects: (**A**) PCA score plot for control and patient samples; (**B**) PCA score plot for control and patients with tumor subtypes; (**C**) PCA loading plot; (**D**) box plots of SM, DG, and TG levels for control and tumor samples.

Supervised orthogonal projections to latent structures discriminant analysis (OPLS-DA) improved differentiation between groups when both sexes were combined, achieving an average accuracy of 74.9 % based on 100-fold cross validation in classifying sample types (**Fig. 3A** and **3B**). The lipid species with the most pronounced concentration differences between tumor subtypes and healthy volunteers were visualized using a volcano plot (**Fig. 3C**) and heat maps (**Fig. 3D**), where lipids downregulated in patients are marked in blue, and those upregulated are shown in red. SM lipid species with an odd carbon number in their fatty acyl chains, such as SM 39:1, SM 41:2, SM 41:1, and SM 33:1, demonstrate the most significant decrease in patient samples, followed by alkyl/alkenyl glycerophospholipids, such as PC O-32:1, PC O-36:4, PC O-38:5, PE O- 38:6, and PE O-38:5. Conversely, several DG species, such as DG 34:3, DG 34:2, DG 34:1, DG 34:0, DG 36:2, and DG 36:1 exhibit the most significant increases in tumor samples, followed by selected TG lipid species (*e.g.*, TG 48:0, TG 51:5, TG 52:1, TG 54:3). The representative lipid species (SM 39:1 and DG 34:1) with the highest concentration differences between the two groups are illustrated in **Fig. 3E** using box plots.

**Figure 3.**
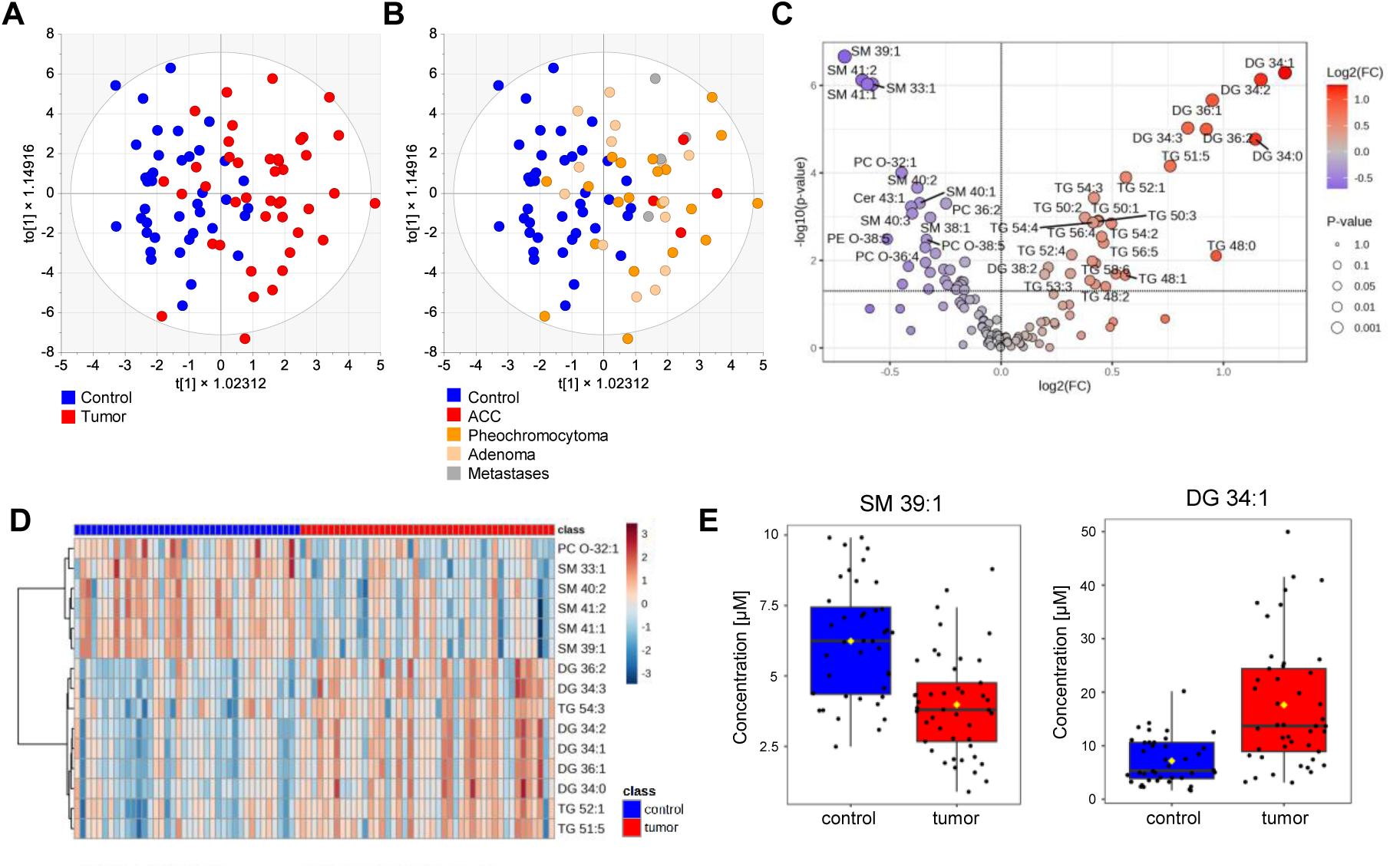
Plots of lipid dataset for both sexes combined: (**A**) OPLS-DA score plot for control and tumor samples; (**B**) OPLS-DA score plot for control and patients with tumor subtypes; (**C**) volcano plot; (**D**) heatmaps showing top 15 lipid species; and (**E**) box plots of lipids showing the largest differences between two groups.

We also evaluated the influence of sex on the results. The group differentiation remained similar when analyzing only female subjects, with an average accuracy of 70.6 %. In contrast, accuracy slightly declined to 69.2%, when only male subjects were included. However, the dysregulation patterns were remarkably consistent across sexes, characterized by a decrease in SM with odd carbon number and the increase in DG with 34 and 36 carbon atoms in patient samples. Corresponding OPLS-DA score plots, volcano plots, heatmaps, and boxplots are shown in Supplementary Material **Fig. S1**. Nevertheless, due to the limited number of subjects, we focused on combining males and females in the models to maximize statistical power by increasing the sample size per group.

We also investigated differences between two groups based on clinical biochemistry and OS parameters. Key findings are visualized in the volcano plot shown in Supplementary Material **Fig. S2**. Patient samples exhibited elevated levels of TG and MDA, while levels of HDL, GSH, and PON1 were reduced compared to healthy subjects.

### Differences Between Healthy Volunteers and Three Types of Adrenal Tumors

As part of our investigation, we compared individual adrenal tumors types to healthy subjects. Our initial focus was on comparing healthy volunteers with individuals diagnosed with confirmed malignant tumors (ACC, metastases), as well as with those diagnosed as benign tumors (adenoma and pheochromocytoma), both of which had presented as benign lesions at their initial diagnosis and confirmed as such by subsequent surgery and histopathological examination. OPLS-DA score plots for both comparisons are shown in **Fig 4**. Better separation was observed between healthy volunteers and malignant tumors (ACC and metastases), as shown in **Fig. 4A**, compared to the separation seen with benign tumors shown in **Fig. 4B**. The lipid species with the most significant differentiation between groups are illustrated in the heatmaps (**Fig. 4C**) and in the volcano plots (**Fig. S3**). The lipidomic profiles showed notable similarities across tumor types, including decreased levels of SM, particularly those with the odd number of carbons (SM 39:1, SM 41:1, and SM 41:2), and increased levels of DG with 34 and 36 carbon atoms, as well as increased levels of TG, such as TG 54:3, TG 54:2, TG 52:1, and TG 51:5. However, some lipidomic profile differences were observed. We found an association of alkyl/alkenyl phospholipids species, such as PC O-36:4, PE O-38:6, and PE O-38:5, with malignant tumors as their concentrations are significantly decreased in tumor patient samples compared to healthy controls and without any significant difference with other two types of adrenal tumors: pheochromocytoma and adenoma. A similar trend was observed for other lipid species, including LPC 20:5, LPC 18:2, PC 38:2, and CE 20:5, which were significantly reduced in malignant tumor samples compared to healthy subjects. This finding was not confirmed in pheochromocytoma and adenoma. Moreover, the predictive power was notably higher for distinguishing healthy subjects from those with malignant tumors (ACC and metastases) than the differentiation with benign tumors (pheochromocytoma and adenoma). ROC curves for malignant and benign tumors are shown in **Fig. 4D** and **4E**, with the malignant group achieving AUC values of 0.933 and an average classification accuracy of 85.5 %, while the group of patient with pheochromocytoma and adenoma showed an AUC of 0.784 and an average accuracy of 71.4 %. The average accuracy was calculated based on 100-fold cross validation. Representative SM and DG species, belonging to the most regulated lipids are visualized using box plots, as shown in **Fig. 4F**.

**Figure 4.**
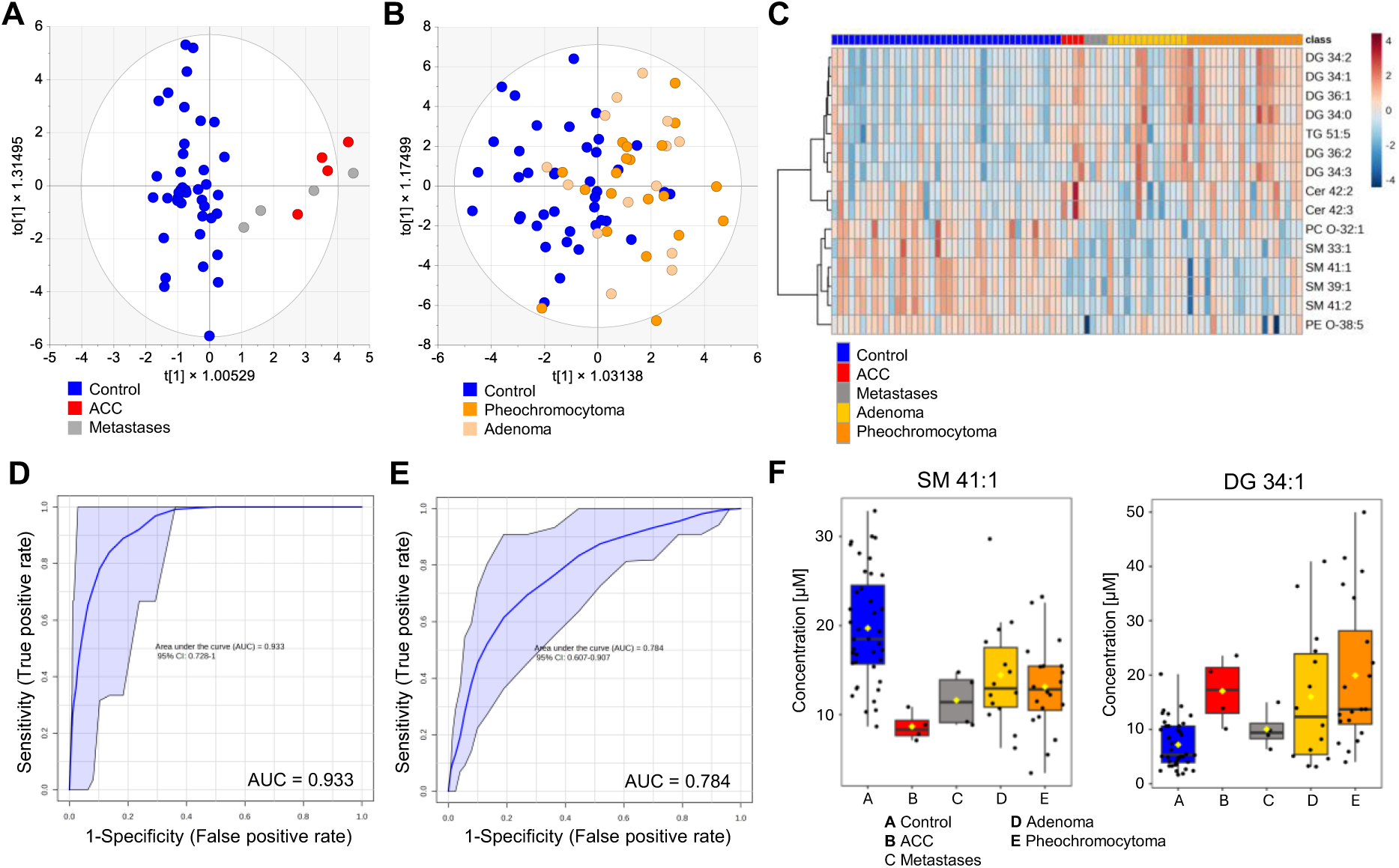
OPLS-DA score plot (lipid dataset) prepared for control samples and: (**A**) malignant tumors (ACC and metastases), (**B**) pheochromocytoma + adenoma tumors; **heatmap** (**C**) showing top 15 lipid species; **ROC curve (lipid dataset) for:** (**D**) malignant tumors, (**E**) benign tumors; and **box plots** (**F**) of representative lipid species.

The benign tumors were further divided into two subtypes: pheochromocytoma and adenoma and compared to healthy volunteers, as shown in **Fig. S4**, due to the potential although infrequent malignant behavior associated with pheochromocytoma. OPLS-DA score plot showed better differentiation for pheochromocytoma than for adenoma samples compared to healthy subjects (**Fig. S4A, S4D**), which is also expressed using ROC plots. ROC curves for pheochromocytoma and adenoma are presented in **Fig. S4B, S4E**, displaying AUC values of 0.800 and the average accuracy 72.7% for pheochromocytoma, and 0.711 and 64.4% for adenoma. The most dysregulated lipid species (SM, DG, and TG) between groups are illustrated in the volcano plots (**Fig. S4C, S4F**). Significant differences are observed in several TG species with increased concentrations in pheochromocytoma compared to healthy volunteer samples (**Fig. S4C**). In contrast, only two TG species show significant differences with increased levels in adenoma tumor patients compared to healthy volunteers (**Fig. S4F**). On the other hand, ceramides with high number of carbons, such as Cer 41:2;O2 and Cer 42:3;O2, showed significant difference (decreased levels) in adenoma tumor samples compared to control samples, neither observed in pheochromocytoma nor in malignant tumors. Due to the limited number of malignant tumors samples, we did not further subdivide them into ACC and metastatic groups, which allowed us to get bigger sample sizes for developing reliable prediction models.

We also applied the same statistical models for the clinical biochemistry and OS parameters. However, we were unable to develop predictive models without overfitting, showing much weaker prediction power for routine clinical biochemistry and OS parameters compared to the lipid dataset in the current set of samples.

### Differences Between Adrenal Tumor Types

Another important part of this project is also to identify the differences between individual adrenal tumor subtypes, which can often be easily misinterpreted. We focused on differences between cortex adrenal tumors, such as malignant ACC and benign adenoma. **Fig. 4F and 5A** show the example of the most regulated lipid species (SM 39:1 and SM 41:1), indicating that the lipid profiling demonstrates similar trends (decreased levels) compared to healthy subjects, but the decrease in concentration is notably more pronounced in ACC than in adenoma. In contrast, ceramides, such as Cer 42:3;O2, Cer 42:2;O2, and Cer 34:1;O2 showed higher levels in primary ACC compared to adenoma samples (**Fig. 5B-C**). These ceramides also showed significant differences between ACC and other tumors, such as pheochromocytoma or metastases, as well. We also compared lipid data between adenoma and pheochromocytoma tumors, but no significant differences were observed. No differences were also observed between adrenal cortex (ACC + adenoma) and medulla (pheochromocytoma) tumors. The clinical biochemistry and OS parameters underwent the same investigation as lipid dataset, but only total thiols levels were significantly decreased in ACC samples in comparison to other adrenal tumors, as shown **in Fig. 5D**.

**Figure 5.**
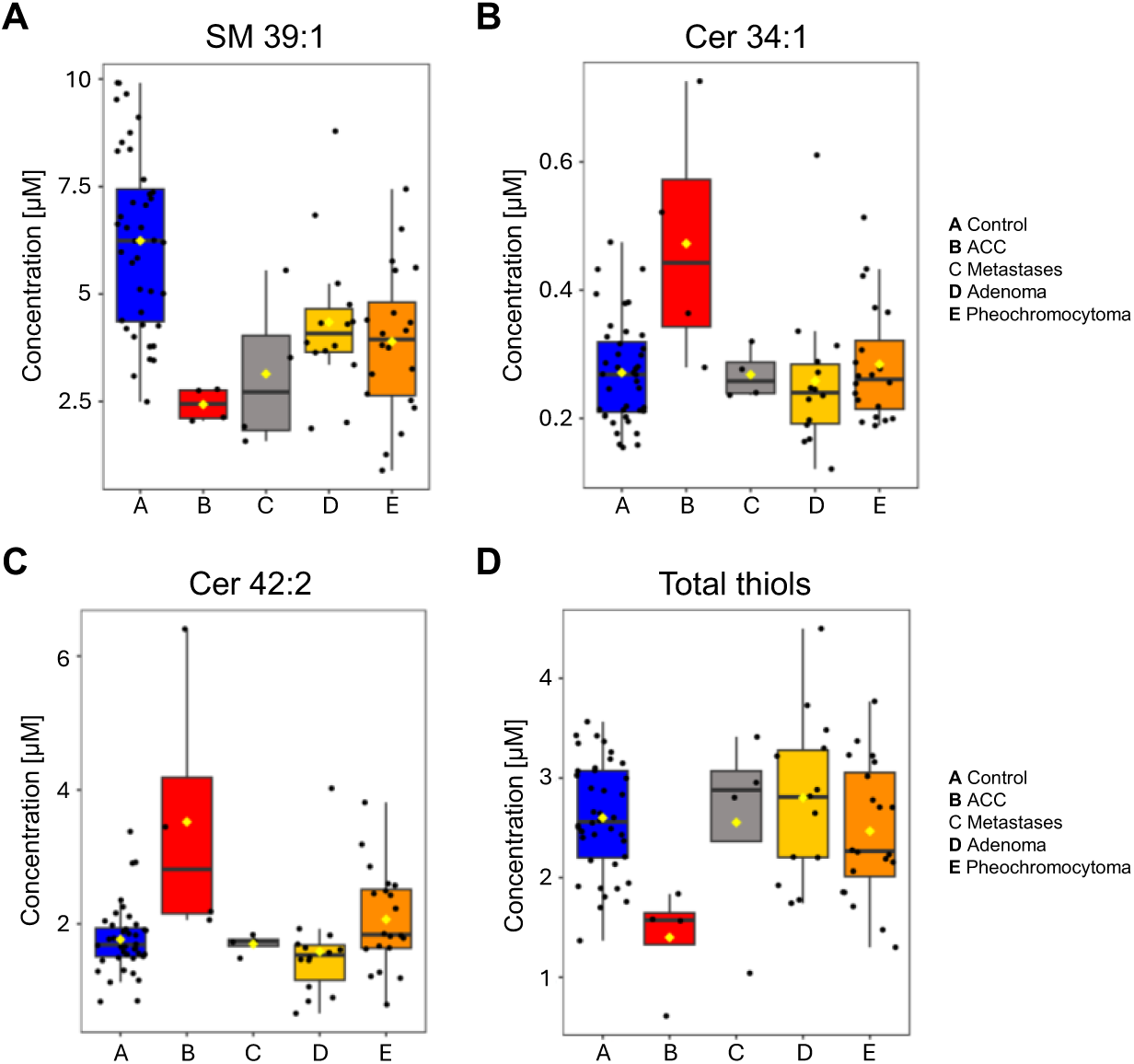
Box plots of representative lipid species, which showed the most significant differences between ACC and other tumors.

### Improvement of Prediction Models

Both the clinical biochemistry/OS parameters and lipid dataset were combined and analyzed to enhance the prediction performance of our statistical models. Only the TG variable from the clinical biochemistry dataset was excluded to prevent overlap with MS data. In the case of malignant tumors, we were able to achieve a higher prediction power for combined datasets, as demonstrated in **Fig. S5A**. The AUC increased from 0.933 to 0.952 and the average accuracy based on 100-fold cross validation increased from 85.5% (lipid dataset only) to 87.2% (clinical biochemistry/OS parameters and lipid dataset combined). On the other hand, for the other tumor types, the combined datasets did not result in higher prediction power, as the AUC and the average accuracy were almost identical for either pheochromocytoma (AUC 0.800 → 0.802; accuracy 72.7% → 73.1%) or adenoma (AUC 0.711 → 0.711; accuracy 64.4% → 66.3%), as shown in **Fig. S5B-C.**

As discussed in the previous section, it is important not only to identify differences between control and individual tumor types but also to differentiate between various types of adrenal tumors, such as malignant primary ACC and benign adenomas. Our objective was to predict the behavior of adenoma samples using a predictive model based on OPLS-DA (clinical biochemistry/OS parameters and lipid dataset combined) that was created for control and malignant tumors. As visualized in **Fig. 6A**, the model predicted that 57% of adenoma samples was classified as control samples, while the remaining cases were classified as uncertain. Notably, none of the adenoma samples were predicted as malignant tumor.

**Figure 6.**
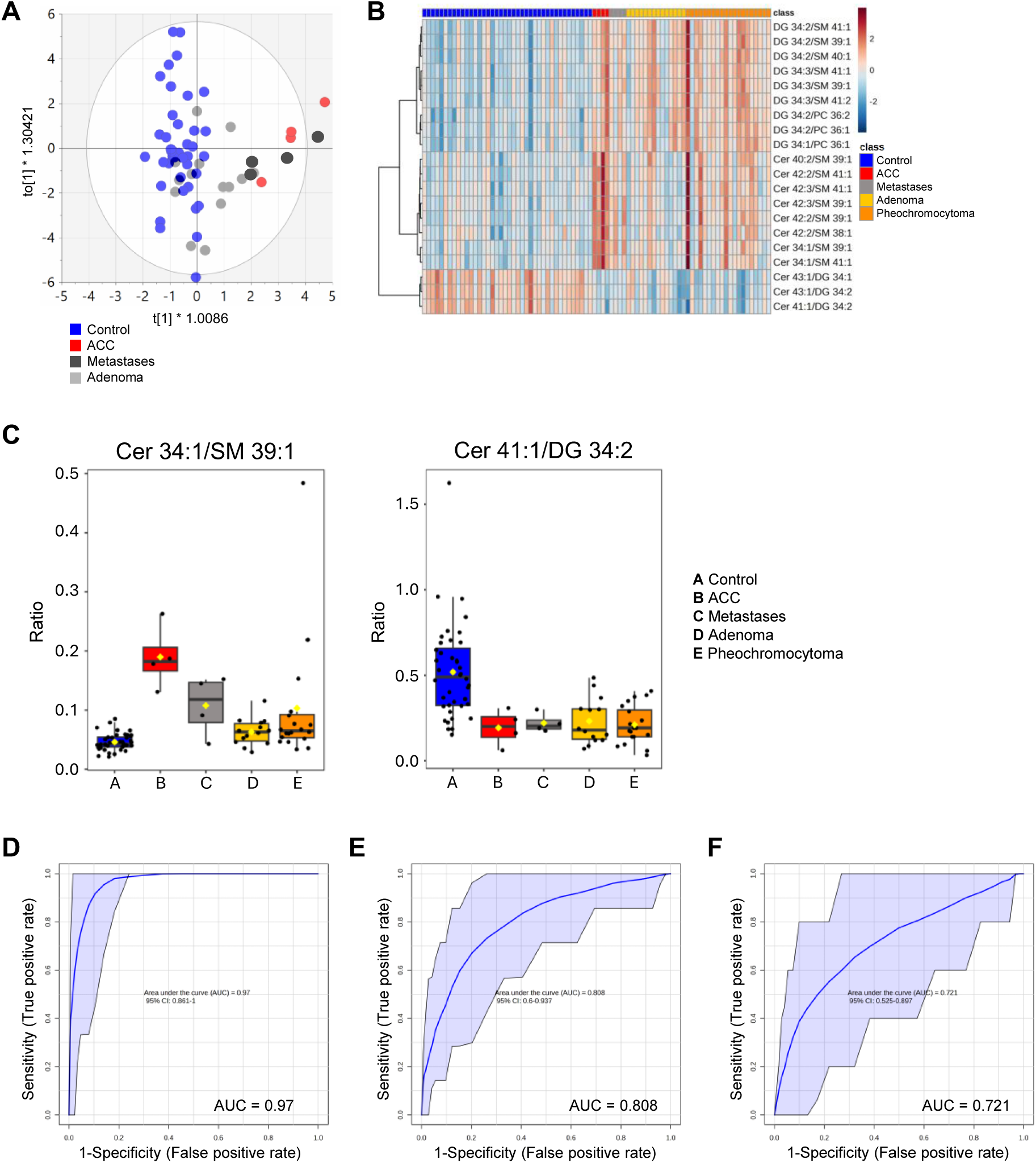
Statistical visualizations: (**A**) OPLS-DA predictive model, (**B**) heatmap of lipid-to-lipid ratios, (**C**) box plots of selected lipid-to-lipid ratios, and (**D**) ROC plots for malignant tumors (ACC and metastases), (**E**) pheochromocytoma, and (**F**) adenoma.

As previously reported [23-25], the lipid-to-lipid ratio provides valuable supporting information and may enhance predictive power. Therefore, we tried to apply this approach to our sample set. The lipid-to-lipid ratios were selected based on their AUC (≥ 0.8) and p-value (≤ 0.05). The top 20 ratios across the individual groups are visualized in a heatmap in **Fig. 6B**. We observed a trend, where the ratios of long chain Cer and DG with 34 and 36 carbon numbers more effectively distinguish control samples from tumor samples, as illustrated in the example (Cer 41:1/DG 34:2) and shown in the box plot in **Fig. 6C**. On the other hand, ratios between Cer and SM with odd carbon number can better distinguish malignant tumors for controls and other adrenal tumors, shown in the example of Cer 34:1/SM 39:1 in **Fig. 6C**. The predictive power of lipid-to-lipid ratios and the individual lipids involved in these ratios is summarized in **Table S2**. As described above, individual lipids generally exhibit lower predictive power compared to their ratios. For malignant tumors, DG 36:2 has AUC = 0.797, PE O-38:5 has AUC = 0.881, while their ratio demonstrates AUC = 0.978, highlighting the enhanced predictive power when the ratio is considered. In this particular case, PE O-38:5 exhibits lower and DG 36:2 exhibits higher levels in malignant tumors compared to controls, therefore, their combination can magnify the differences between these two groups.

For the next step, we used the lipid-to-lipid ratio as an input dataset to develop our prediction models. The input dataset included both clinical biochemistry/OS parameters and lipid dataset, but individual lipids involved in the lipid-to-lipid ratios were excluded to avoid potential overfitting. All datasets used for the improved statistical models are detailed in **Table S3-5** in the Supplementary Material. When the lipid-to-lipid ratio was incorporated, we observed models with higher predictive power, particularly in the case of malignant tumors, as shown in **Fig. 6D** with the ROC plot. The AUC increased to 0.97, with the average accuracy of 89.3% based on 100-fold cross-validation. Prediction power was also slightly improved for pheochromocytoma (AUC 0.808, accuracy 73.9%) and adenoma (AUC 0.721, accuracy 67.3%). However, this approach should be applied with caution, and it is essential to validate these results using a larger, independent sample set. Nonetheless, the lipid-to-lipid ratio shows promise as a tool to enhance the power of statistical models and warrants further exploration.

### Time Points Collection

We further examined multiple collection time points (pheochromocytoma and adenoma) to monitor changes in lipid profiles after surgery (**Table S6**). As illustrated in **Fig. S6A**, PCA score plot did not reveal distinct clustering patterns for samples collected after the initial time point. However, we specifically focused on clinical biochemistry/OS parameters that demonstrated significant differences between tumor samples and controls, evaluating whether their levels changed over time (after surgery). **Fig. S6B-E** presents box plots for key markers including GSH, HDL, MDA, and TG. HDL levels, which initially decreased in tumor samples, remained consistently low at all time points. GSH exhibited concentrations that decreased further at 1.5 months after surgery before returning (12-30 months after surgery) to levels comparable to the initial time points (day of surgery). In contrast, TG and MDA showed different temporal patterns. These markers displayed no significant differences between the initial and 1.5 months after surgery. However, their concentrations had returned to levels comparable to control samples by the third collection (12-30 months after surgery).

We subjected the lipidomic dataset to identical statistical procedures. SM with odd-numbered carbon chains, which exhibited reduced levels in tumor samples, remained consistent across all study (time points) without temporal variations. In contrast, DG followed patterns similar to TG and MDA. Specifically, DG showed no significant concentration changes between the initial and 1.5 months after surgery (second collection). Their levels were normalized to match control samples by the third collection. PC demonstrated yet another pattern, with concentrations progressively decreasing at both the 1.5-month and 12-30 month time points after surgery. Box plots of representative lipid species in **Fig. S6F-H** illustrate these trends.

## DISCUSSION

This pilot study demonstrates that lipidomic profiling of human serum can identify distinct signatures associated with various subtypes of adrenal tumors. The analysis was conducted on 42 patients diagnosed with various adrenal tumor subtypes and metastases, along with 40 carefully matched controls. The control group was selected to closely match the patients in terms of age, BMI, and sex, minimizing potential discrepancies in lipid concentrations due to anthropometric differences [26, 27].

The most dysregulated lipid species between control and tumor samples belonged to the SM, DG, and TG lipid classes. In particular, SM with odd-numbered carbon chains (SM 39:1, SM 41:1, SM 41:2) decreased in patients, and DG with 34 and 36 carbon chains showed an opposite effect, elevated levels in patients. These findings are consistent with the notion that lipid metabolism is closely related to tumorigenesis, with specific lipid species contributing to membrane dynamics, energy balance, and oncogenic signaling pathways [12–14]. SM and DG are known to play important roles in cell proliferation and apoptosis. Specifically, imbalances in SM may reflect heightened sphingomyelinase activity or altered biosynthesis, processes linked to oncogenic signaling. Meanwhile, DG serves as a lipid second messenger promoting activation of protein kinase C and other pathways that drive tumor cell growth and survival. More broadly, aberrant lipid metabolism, including glycerolipid remodeling, has emerged as a hallmark of cancer biology. Reduced levels of odd-chain SM may reflect alterations in membrane composition or increased degradation, whereas elevated DG levels suggest enhanced lipolytic activity or de novo synthesis associated with oncogenic transformation [28-35]. Therefore, SM and DG may serve as potential markers for distinguishing between healthy subjects and those with adrenal tumors.

The most pronounced lipid alterations were observed in malignant tumors (ACC, metastases), followed by pheochromocytoma and adenoma. Our findings suggest that lipidomic analysis has potential as a valuable tool for distinguishing malignant ACC and metastases from other adrenal tumors. Notably, we identified a specific association between malignant tumors and decreased levels of alkyl/alkenyl glycerophospholipids and increased levels of long-chain Cer compared to pheochromocytoma and adenoma. These lipid species may serve as promising markers for differentiating ACC from other adrenal tumor subtypes.

The clinical biochemistry/OS parameters underwent the same evaluation. There are several markers that can indicate increased OS in the body and can be associated with cancer, such as adrenal tumors. Reduced levels of plasma total thiols, PON1 activity in plasma, GSH in erythrocytes, SOD, GPx, and catalase activity in erythrocytes may indicate increased OS in the body. On the other hand, increased levels of markers, such as MPO activity in plasma, MDA in plasma (LPX) and MDA in erythrocytes, and GR in erythrocytes may indicate increased OS in the body. However, these parameters exhibited inferior discriminatory power in identifying the subtypes of adrenal tumors. Although some OS parameters, such as MDA and GSH showed trends consistent with previous reports [18, 30], their variability and lower predictive accuracy limit their diagnostic utility in this context. On the other hand, combining clinical biochemistry/OS parameters with lipidomic dataset modestly improved model performance, especially for malignant tumors.

Importantly, the implementation of lipid-to-lipid ratios significantly enhanced the predictive performance of statistical models. This approach magnifies subtle differences between groups by integrating biological relationships between lipid species. In particular, ratios involving Cer and DG or SM showed the highest discriminatory capacity, highlighting their potential as composite markers.

Time point results showed that TG, DG, and MDA would appear suitable for monitoring tumor response during and after treatment interventions (adrenalectomy). In contrast, GSH and HDL can work more as the supportive information to differentiate control and tumor samples.

Despite these promising results, this study is limited by its relatively small sample size and retrospective design. While initial findings are encouraging, larger prospective cohorts are required to validate the diagnostic value and robustness of these lipidomic signatures. Additionally, further mechanistic studies are warranted to understand the biological relevance of specific lipid alterations in adrenal tumorigenesis.

## METHODS

### Chemicals and Reagents

LC-MS grade solutions and additives, such as acetonitrile, methanol, 2-propanol, hexane, ammonium carbonate, and ammonium acetate (Honeywell, Riedel-de Haën, Germany), were purchased from Thermo Fisher Scientific (Waltham, MA, USA). Chloroform (Lichrosolv) was purchased from Merck (Darmstadt, Germany) and supercritical carbon dioxide (scCO_2_, 4.5 grade purity) from Messer (Bad Soden, Germany). Deionized water was obtained using a Milli-Q water purification system (Millipore, Molsheim, France). Lipid class internal standards, such as phosphatidylcholine 15:0/18:1-D7 (PC 33:1 D7), phosphatidylcholine 18:1-D7 (LPC 18:1 D7), phosphatidylethanolamine 15:0/18:1-D7 (PE 33:1 D7), sphingomyelin 18:1/18:1-D9;02 (SM 36:2 D9), triacylglycerol 15:0/18:1-D7/15:0 (TG 48:1 D7), diacylglycerol 15:0/18:1-D7 (DG 33:1 D7), monoacylglycerol 18:1-D7 (MG 18:1 D7), cholesteryl ester 16:0-D7 (CE 16:0 D7), ceramide 18:1-D7/18:0;O2 (Cer 36:1;O2 D7), and cholesterol-D7 (Chol-D7) manufactured by Avanti Polar Lipids (Alabaster, AL, USA) and supplied by Merck.

### Clinical Study

The present clinical study was conducted from 2016 to 2022 in University Hospital in Olomouc at its three departments: Department of Radiology, Department of Urology, and Third Department of Internal Medicine - Nephrology, Rheumatology and Endocrinology. The clinical study was approved by the Ethics Committee of the University Hospital and the Faculty of Medicine and Dentistry, Palacký University Olomouc, Czech Republic (reference number: 121/16). All study participants signed an informed consent.

The inclusion criteria for all patients (N=42; 21 females and 21 males) required to undergo adrenalectomy, have comprehensive CT imaging documentation stored in the picture archiving communicating system (PACS), and receive definitive histopathological diagnosis of adrenal tumor. Blood was collected from each patient on the day of surgery. To evaluate serum lipidomics stability, samples were collected 1.5 (10, female and 16, male) and/or between 12-30 months (8, female and 11, male) after surgery, respectively. In 14 patients (5, female and 9, male) blood samples were collected at all time points. Blood samples from healthy volunteers were collected only once (during the course of clinical trial). Blood was used for serum and plasma preparation and the aliquots (lipidomic analysis, markers, and selected parameters of clinical biochemistry and OS) were stored at – 80°C before the analysis.

The inclusion criteria for all healthy volunteers (N=40; 21, female and 19, male) required morphologically normal adrenal gland. All volunteers underwent MRI of the abdomen: 1) to detect any enlargement of adrenal glands, 2) to identify any tumors in the peritoneal cavity or retroperitoneum, and 3) to assess for other serious pathological conditions, particularly acute inflammatory and malignant changes. The MRI examination was performed using a 1.5 T Siemens machine, employing fast gradient sequences in both axial and coronal planes. Another inclusion criterion was a documented history free of malignancy, systemic or other chronic diseases, together with the absence of clinical signs/symptoms of acute illnesses, such as infections or trauma. Additionally, healthy volunteers in the control group were prohibited from taking short- and long-term medications.

Clinical and other characteristics of all participants including age, BMI, sex, routine clinical biochemistry parameters (total cholesterol, HDL, LDL, glucose, and triacylglycerols (TG)) were measured spectrophotometrically using the Cobas 8000 analyzer (Hitachi). Furthermore, in each participant, selected OS parameters, such as plasma total thiols (total SH), plasma malondialdehyde (LPX), myeloperoxidase (MPO) and paraoxonase 1 (PON1) activity in plasma, malondialdehyde (MDA) levels in erythrocytes, glutathione (GSH) levels in erythrocytes, superoxide dismutase (SOD) activity in erythrocytes, glutathione peroxidase (GPx) activity in erythrocytes, glutathione reductase (GR) activity in erythrocytes, and catalase (CAT) activity in erythrocytes were measured. Blood OS parameters were measured following the methodology described in [36], with the exception of PON1 activity, which was estimated according to the protocol described in [37].

### Lipid Extraction

Twenty-five µL of sample, 20 µL of internal standard mixture (**Table S7 in Supplementary Material**), and 3 mL of chloroform/methanol (2:1, *v/v*) were added together and sonicated (15 min, 40 °C). After sonication, 600 µL of aqueous ammonium carbonate (250 mM) was added, then sonicated again (15 min, 40 °C) and centrifuged (3000 rpm, 3 min). The organic (lower) phase was transferred to a new vial. Additional 2 mL of chloroform was added to the aqueous phase and followed by sonication (15 min, 40 °C) and centrifugation (3000 rpm, 3 min). The organic phase was removed and combined with the previous one. The collected organic phase was evaporated under the nitrogen flow and then dissolved in 500 µL of chloroform/methanol (1:1, *v/v*) mixture. After a 5-fold dilution with chloroform/methanol (1:1, *v/v*), the sample was injected into ultrahigh-performance supercritical fluid chromatography – mass spectrometry (UHPSFC/MS) system for the analysis.

### UHPSFC/MS Conditions

Lipid separation using UHPSFC (Acquity UPC^2^ instrument from Waters; Milford, MA, USA) was performed on column Viridis BEH (100×3mm, 1.7 µm) with the following conditions: the column temperature 60 °C, the flow rate 1.9 mL/min, and the injection volume 1 µL. The injection needle was washed with hexane/2-propanol/water (2:2:1, *v/v/v*) after each injection. The following linear gradient was performed using scCO_2_ and methanol (30 mM ammonium acetate + 1% of water) used as a modifier: 0 min – 1% modifier, 1.5 min – 16% modifier, 4 min – 51% modifier, 7 min – 51% modifier, 7.51 min – 1% modifier, and the equilibration with the total run time of 8 min. The automatic back-pressure regulator was set to 1800 psi and the autosampler temperature to 4°C. Methanol with 30 mM ammonium acetate and 1% of water was used as the make-up solvent with a flow rate of 0.25 mL/min. The UHPSFC was connected with the hybrid quadrupole - time of flight (QTOF) mass spectrometer Synapt G2-Si from Waters with following conditions: sensitivity mode applying positive ESI mode, the mass range of *m/z* 150-1200, the capillary voltage of 3 kV, the sampling cone of 20 V, the source offset of 90 V, the source temperature of 150 °C, the desolvation temperature of 500 °C, the cone gas flow of 50 L/h, the desolvation gas flow of 1000 L/h, and the nebulizer gas flow of 4 bar. Mass spectra were acquired in the continuum mode with scan time 0.5 s and the peptide leucine enkephalin as the lock mass.

### Data Processing

All UHPSFC/MS-QTOF spectra were acquired using MassLynx and processed as follows: noise reduction using the Compression tool followed by the lock mass correction and the conversion from continuum to centroid mode using the MassLynx tool (Accurate mass measure). Retention time windows of individual lipid classes were set to the methods created to obtain intensities (threshold of intensity 3000) using the MarkerLynx tool. Furthermore, these methods for each lipid class were applied to a sequence of samples, which results in a sum table of all *m/z* with the corresponding intensity for each sample, exported as *.txt* file and used for lipid identification, calculation of concentrations, and isotopic correction Type II and Type I using LipidQuant 2.1 [38]. All lipid intensities were normalized to the respective internal standard of the lipid class internal standard and reported as molar concentrations (µM). The data were further analyzed using statistical tools, such as Metaboanalyst and SIMCA. The lipid concentrations were logarithmically transformed (base 10) and Pareto scaled.

### Quality Control of Lipidomic Analysis

Lipids selected for statistical evaluation underwent quality control (QC) process, which included the following criteria: a coefficient of variation (CV) of QC samples below 25%, sample-to-blank ratio (S/B) calculated as the ratio of the average of QC samples and blank samples exceeding 5, and linearity of dilution series assessed using the Pearson correlation coefficient, with the value greater than 0.9 (R²). Lipids that did not meet these QC criteria were excluded from the statistical analysis. Ultimately, 125 lipids across 9 lipid classes were identified and quantified in plasma/serum samples that successfully passed the QC assessment and were subsequently used for statistical evaluation (**Table S8**). The lipid coverage was as follows: CE (n=7), TG (n=44), DG (n=8), ST (n=1), Cer (n=11), SM (n=15), PC (n=22), LPC (n=14), and PE (n=3). **Fig. S7A** displays the PCA score plot, with the QC sample highlighted showing a clustering, demonstrating the high precision of the measurements. As a QC measure, we also analyzed NIST Standard Reference Material (SRM) 1950 Metabolites in Frozen Human Plasma alongside our samples. The lipid concentrations are reported in **Table S9**. This dataset can serve as a valuable resource for the lipidomics community to standardize experiments across different laboratories.

We also assessed potential differences between plasma and serum samples, which were collected from each patient simultaneously. The correlation between these two biological materials was excellent with R^2^ equal to 0.9992 (**Fig. S7B**). However, since some plasma samples were collected using EDTA as the anticoagulant while others utilized heparin, we decided to focus primarily on human serum samples to maintain consistency throughout our analysis. As a result, the following data are presented for human serum samples.

## Abbreviations

ACC: adrenocortical carcinoma
AUC: area under the curve
BMI: body mass index
CE: cholesteryl ester
Cer: ceramide
CT: computed tomography
DG: diacylglycerol
GSH: glutathione
HDL: high-density lipoprotein
LDL: low-density lipoprotein
LPC: lysophosphatidylcholine
MRI: magnetic resonance imaging
OPLS-DA: orthogonal projections to latent structures discriminant analysis
OS: oxidative stress
PC: phosphatidylcholine
PCA: principal component analysis
PE: phosphatidylethanolamine
PET: positron emission tomography
QC: quality control
QTOF: quadrupole time-of-flight
ROC: receiver operating characteristic
SM: sphingomyelin
TG: triacylglycerol
UHPSFC-MS: ultrahigh-performance supercritical fluid chromatography – mass spectrometry.

## Author Contributions

Michaela Chocholouskova: conceptualization, measurements, data curation, visualization, and writing of the original draft; Filip Čtvrtlík: sample and clinical data collection; Zbyněk Tüdös: sample and clinical data collection; Igor Hartmann: sample and clinical data collection; Jan Schovánek: sample and clinical data collection; Jitka Vostálová: biochemical analysis; Jitka Prošková: biochemical analysis; Karel Pacák: consultation; Michal Holčapek: project design, administration, and funding acquisition. All authors participated in the review and editing of the manuscript and approved the final version.

## Data Availability

Data are available at the Department of Analytical Chemistry, Faculty of Chemical Technology, University of Pardubice, on reasonable request from the corresponding author.

