## Supplementary File_figures for "A Pilot Study on Serum Lipidomic Alterations in Patients with Adrenal Tumors"

### SUPPLEMENTARY MATERIAL

**Figure S1. Plots for female: (A)** OPLS-DA score plot; **(B)** OPLS-DA score plot for control and patients with tumor subtypes; **(C)** volcano plot; **(D)** box plots of representative lipids showing the highest differences between the two groups; **(E)** heatmaps showing top 15 lipid species; **Plots for male: (F)** OPLS-DA score plot; **(G)** OPLS-DA score plot for control and patients with tumor subtypes; **(H)** volcano plot; **(I)** box plots of representative lipids showing the highest differences between the two groups; **(J)** heatmaps showing top 15 lipid species.

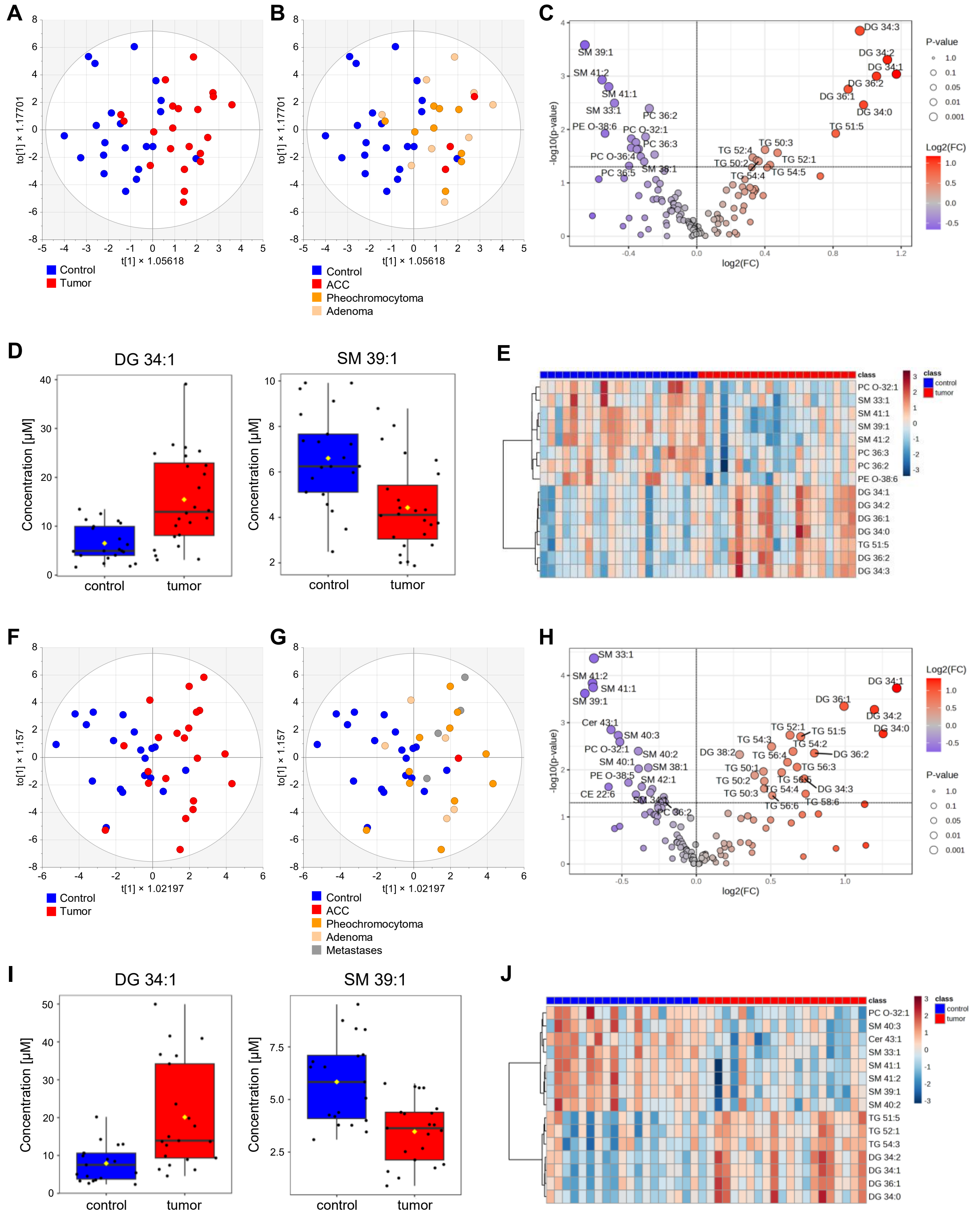

**Figure S2. Selected markers of oxidative stress and routine laboratory lipid dataset, both sexes combined:** (**A**) Volcano plot, (**B**) box plots of TG, (**C**) box plots of MDA, (**D**) box plots of HDL, (**E**) box plots of GSH, and (**F**) box plots of PON1.

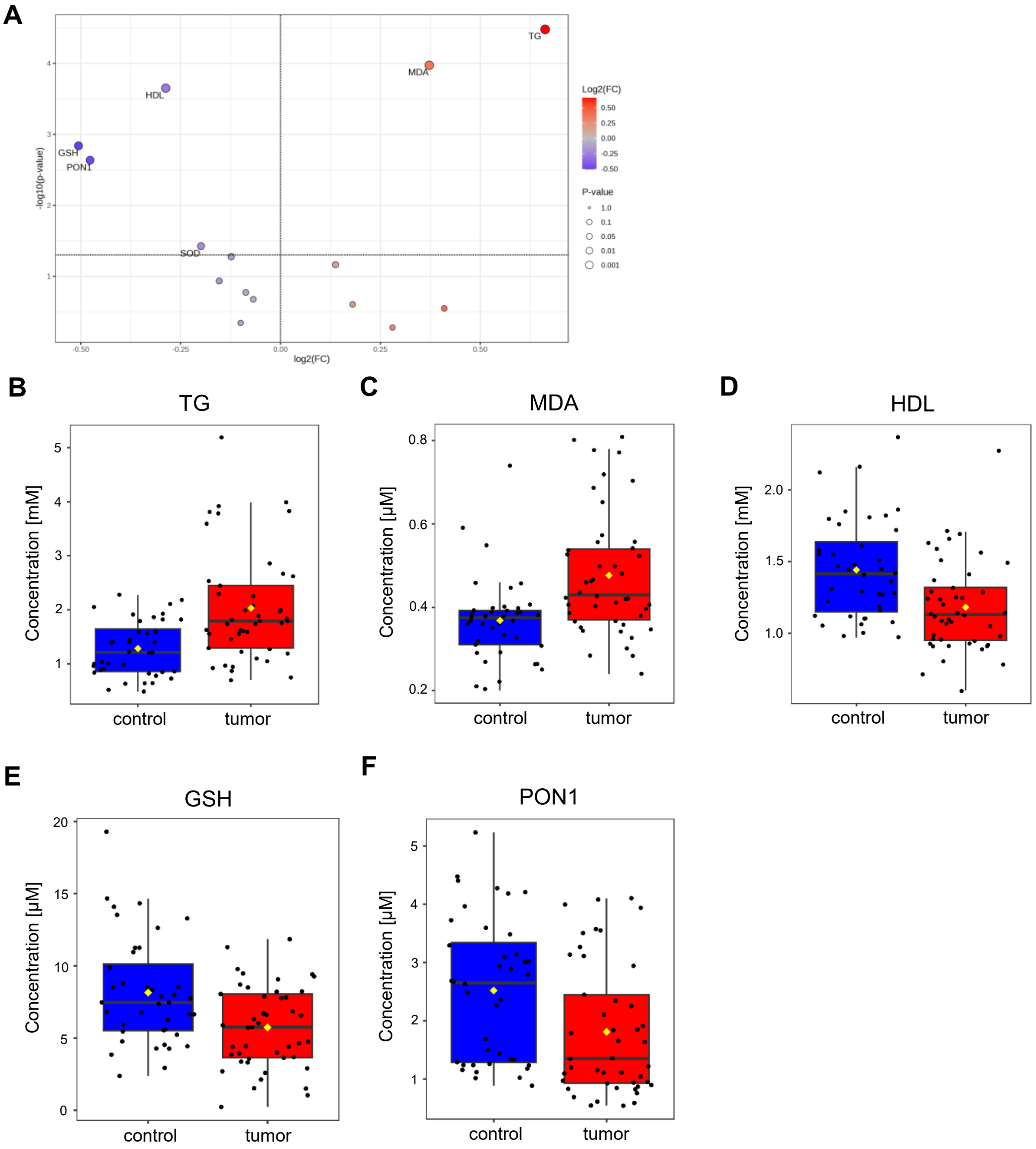

**Figure S3. Volcano plots** created for control samples and: **(A)** malign tumors (ACC and metastases); **(B)** pheochromocytoma + adenoma tumors. Blue color: decreased levels in tumor samples, red color: increased levels in tumor samples.

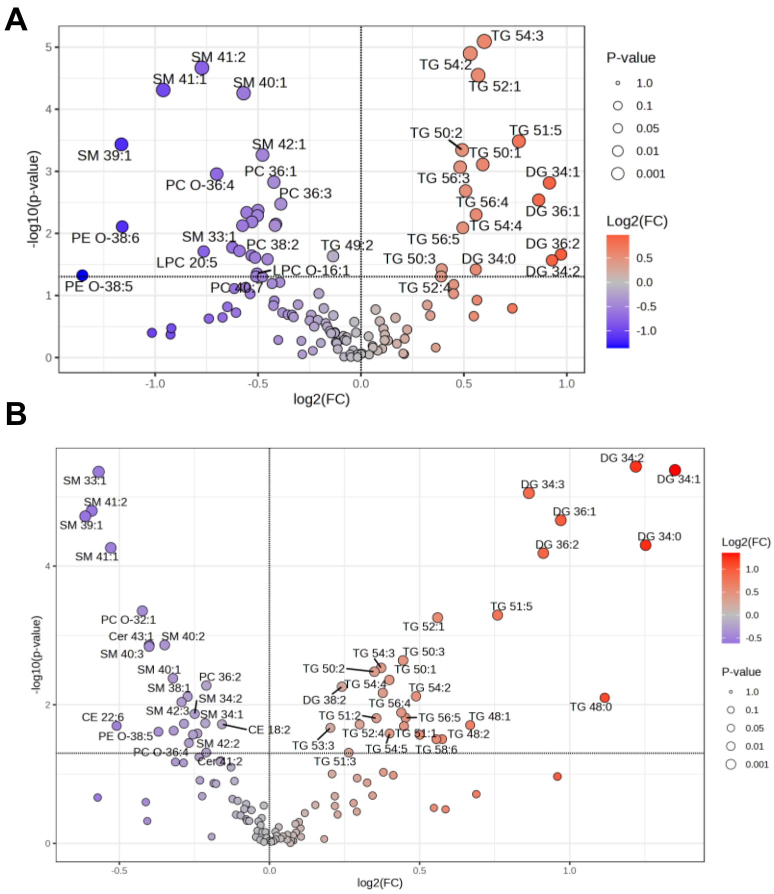

**Figure S4. OPLS-DA score plot created for controls and**: (**A**) pheochromocytoma, and (**D**) benign adenoma; **ROC curve:** (**B**) pheochromocytoma, and (**E**) benign adenoma; and **Volcano plots**: (**C**) pheochromocytoma, and (**F**) benign adenoma.

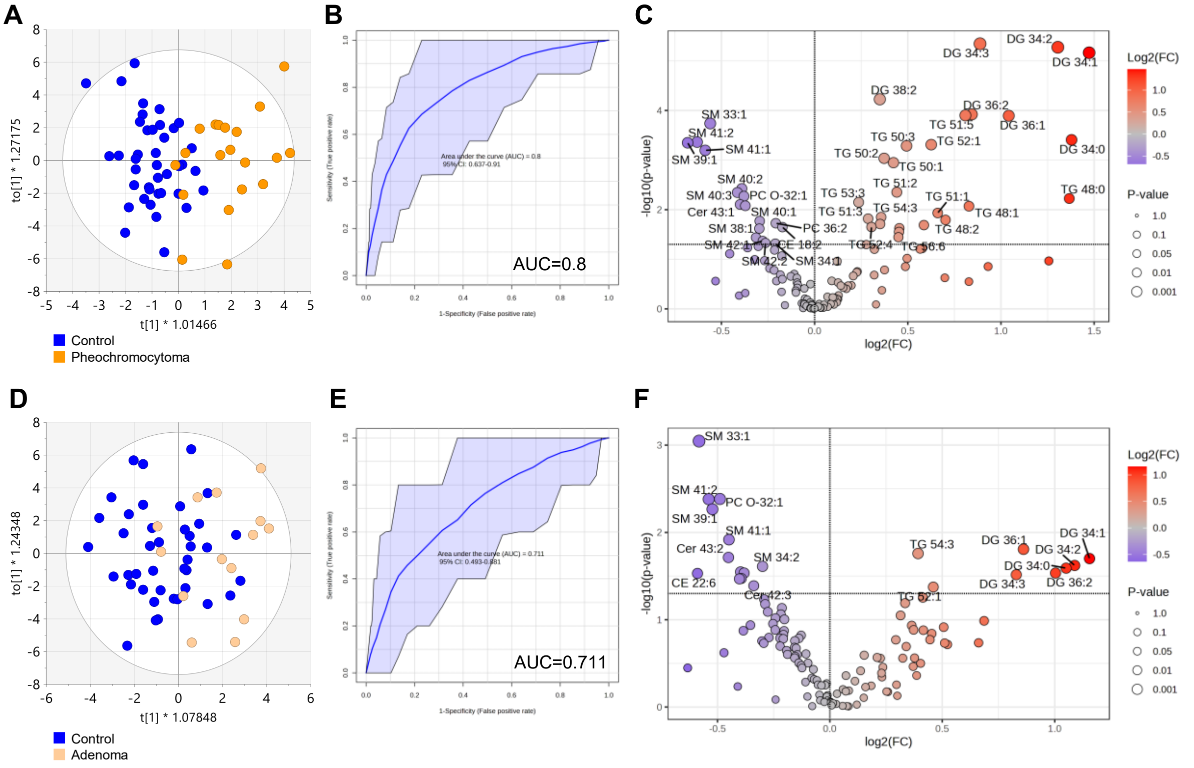

**Figure S5. OPLS-DA score plots** and **ROC plots for: (A)** primary adrenocortical carcinoma/metastases; **(B)** pheochromocytoma; and **(C)** adenoma created for combined clinical biochemistry/OS parameters and lipid datasets.

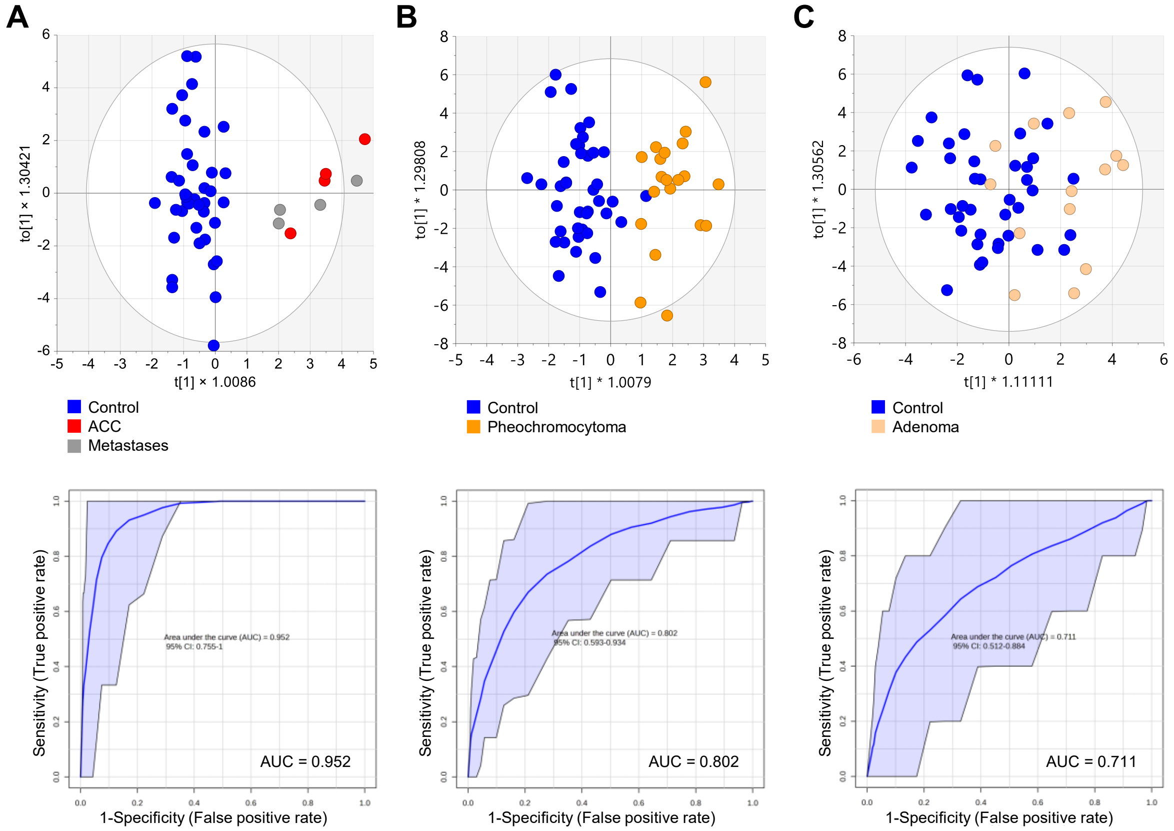

**Figure S6. (A)** PCA score plot comparing controls, tumor samples, and their respective time points; **(B)** GSH distribution represented as box plot**; (C)** HDL distribution represented as box plot; **(D)** MDA distribution represented as box plot; **(E)** TG distribution represented as box plot; **(F)** SM 39:1 distribution represented as box plot; **(G)** DG 34:1 distribution represented as box plot; and **(H)** PC 38:6 distribution represented as box plot.

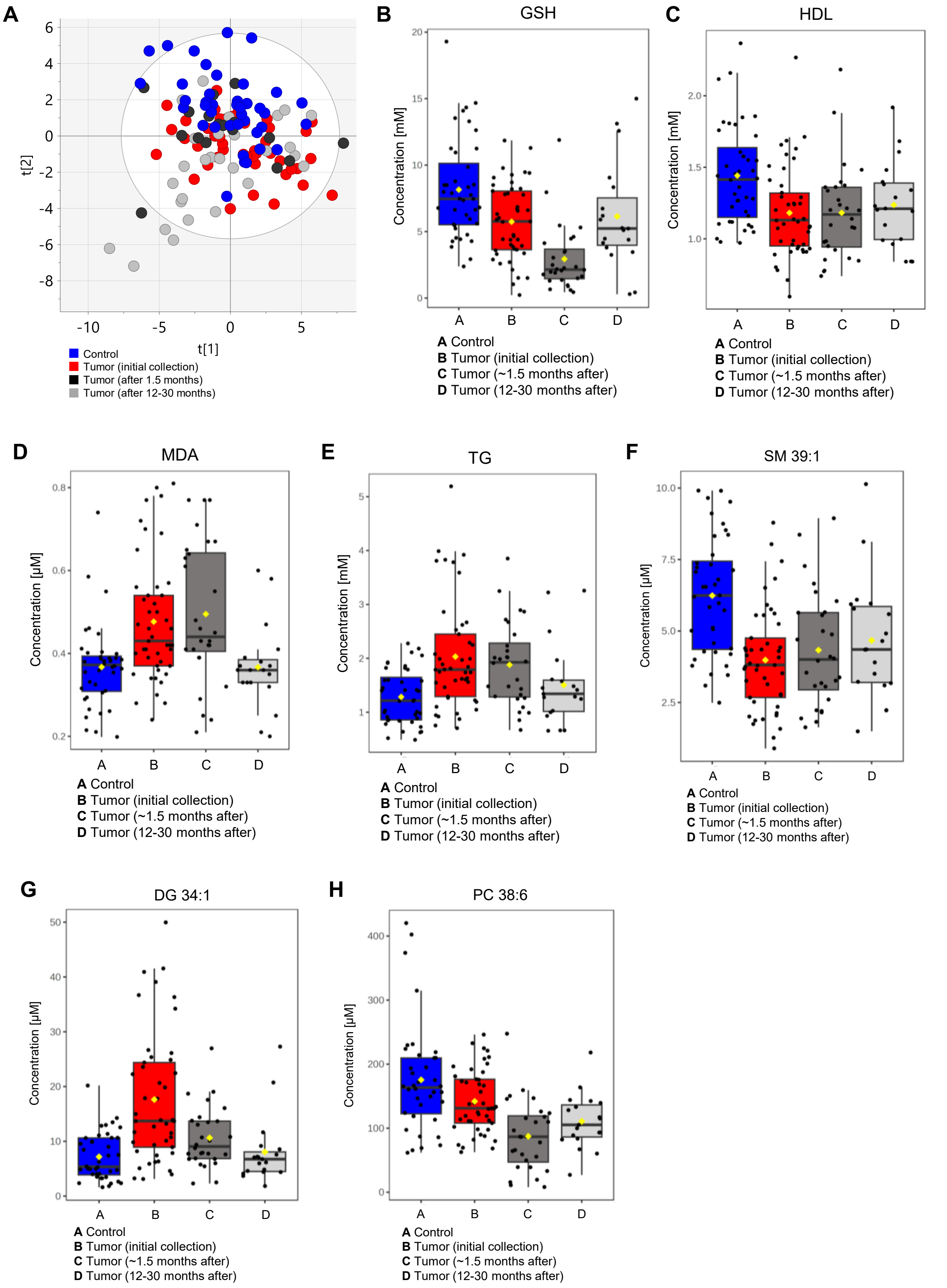

**Figure S7. Quality control: (A)** PCA score plot with clustered and highlighted QC samples: blue - BQC plasma; yellow - BQC serum; green - TQC; and grey – samples. (**B**) Correlation plot for serum and plasma samples.

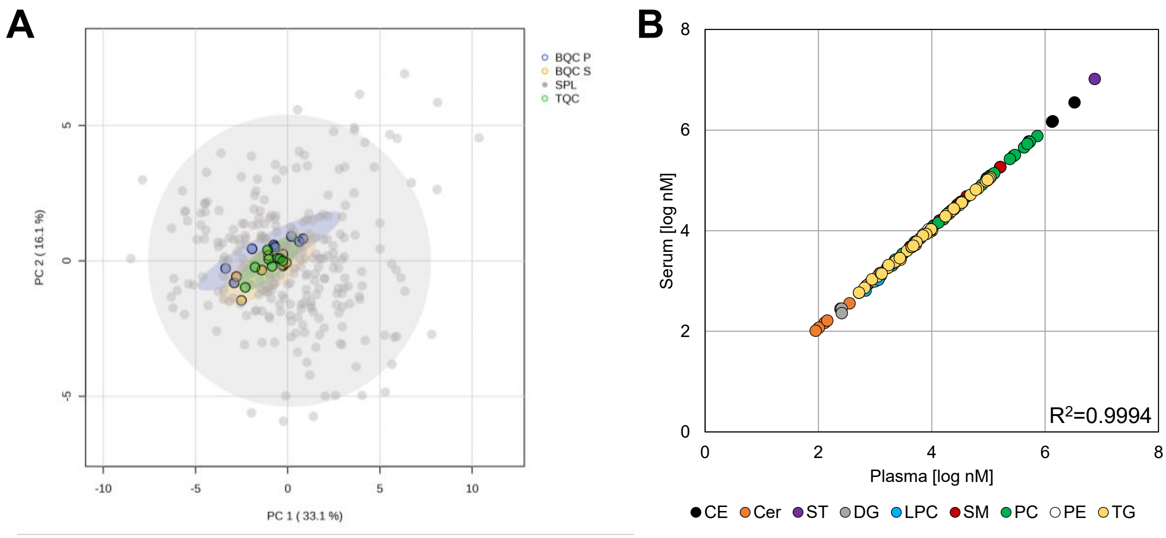
